# Jump-Drop Adjusted Prediction of Cumulative Infected Cases using the Modified SIS Model

**DOI:** 10.1101/2022.07.13.22277574

**Authors:** Rashi Mohta, Sravya Prathapani, Palash Ghosh

## Abstract

Accurate prediction of cumulative COVID-19 infected cases is essential for effectively managing the limited healthcare resources, specifically in India. Historically, epidemiological models have helped control such epidemics. Those models need accurate past data to predict for future. However, for various reasons, we observe sudden drops and jumps in the number of daily reported COVID-19 infected cases on some days, not aligned with the overall trend. If we incorporate those observations in the training data, the model’s prediction accuracy may worsen, as they do not capture the correct trend in the training data. But it is not straightforward for the existing epidemiological models to decide a specific day as a sudden drop or jump. We propose an algorithm that automatically determines any drop or jump days in this work. Then, based on the overall trend in the data, we adjust the number of daily infected cases on those days and decide on the training data based on the adjusted observations. We have applied the proposed algorithm in a recently proposed modified Susceptible-Infected-Susceptible (SIS) to show that adjusted training data gives better prediction accuracy when jump and drop exist in the training data.

## 1 Introduction

Researchers have used different epidemiological models to understand the dynamics of the Coronavirus disease 2019 (COVID-19) since it was first reported in Wuhan city of China [1]. The main focuses of those models are prediction [2], estimation of the basic reproduction number (*R*_0_) [3], trend detection [4], reinfection [5], the effect of preventive measures such as lockdown [6], social distancing [7], etc. All these approaches need high-quality training data to develop meaningful and effective models. We have observed several waves of COVID-19 at distinct points of time in the different parts of the world with different patterns during the last two and half years. Due to many variants of the COVID-19 virus, it may not be wise to consider the entire history data to estimate model parameters. However, there are not many attempts to check whether the available training data is appropriate for model building. [8] proposed an algorithm that optimally decides the length of the training period to predict the number of cumulative infected cases using a modified susceptible infected susceptible (SIS) model.

The daily number of reported COVID-19 infected cases may have high fluctuations due to less testing during festivals (drops), sudden movement of a group (student/workers) people (jumps), etc. These *false* jumps or drops are not aligned with the actual trend of the reported infected cases. On the contrary, we also observe genuine decline (drop) or increase (jump) in the reported infected cases due to the actual trend of COVID-19 spread. This phenomenon is quite common in a large country like India, with a highly dense population in cities and diversity in terms of climate and behavior of people. It is essential to distinguish the difference between the actual trends and false drops/jumps in the reported infected cases. A few false drops and jumps in the training data may result in the poor estimation of model parameters and subsequently dismal prediction performance.

Figure 1 shows daily COVID-19 infected cases from 14 March 2020 to 4 June 2021 in Delhi, the capital city of India. We observe many jumps and drops in the number of infected cases from the overall trend of the curve. There is a visible sharp drop from 7750 to 3330 infected cases on 14 November 2020. On this day (in 2020), Diwali, a major Indian festival, was celebrated. A possible explanation for this drop would be that people with minor symptoms did not get tested on Diwali, or some testing centers were closed or operated only for a few hours. Therefore, this drop can be considered a false drop that is not aligned with the actual trend of the daily infected curve. Such events are usually one-time occurrences and seldom have any impact after they are done. However, if we consider these false drops (or jumps) into the training data, they have the potential to drag the fitted curve towards them, ignoring the actual overall trend. Therefore, it is essential to adjust these drops and jumps to eliminate the impact of such events in the development of models. In other words, these drops and jumps are similar to the notion of outliers or influential points, which may alter the model’s prediction performances. But it is not straightforward for the existing epidemiological models to decide a specific day as a sudden drop or jump in the training data.

**Figure 1:**
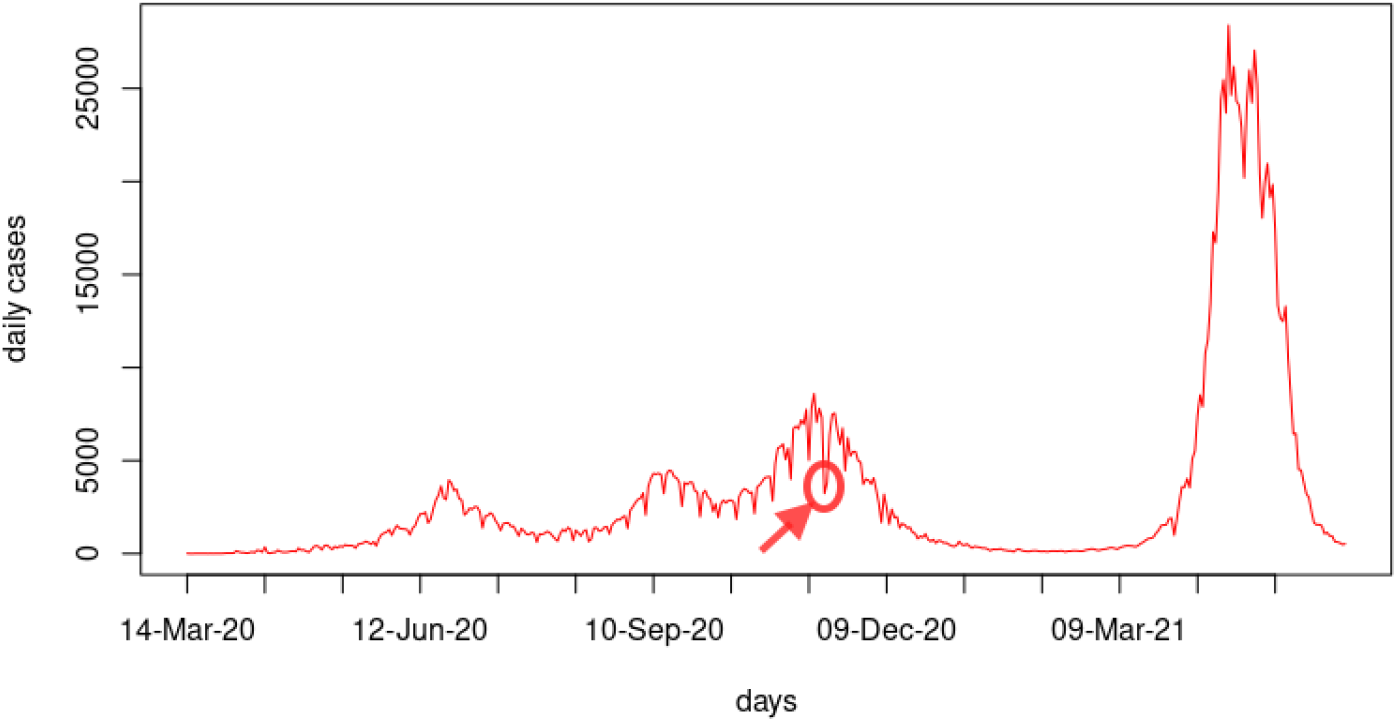
Daily infected COVID-19 cases from 14 March 2020 to 4 June 2021 in Delhi highlighting the drop on 14 November 2020.

In this work, we propose an algorithm that automatically determines any drop or jump day in the training data. Then, based on the overall trend in the training data, we adjust the number of daily infected cases on those days. The effective training data becomes the adjusted daily COVID-19 infected cases. To show that the adjusted training data gives better prediction accuracy when jump and drop exist in the training data, we have applied the proposed algorithm in a recently proposed modified Susceptible-Infected-Susceptible (SIS) [8]. However, this algorithm is useful for any model to clean the training data for better prediction accuracy. Note that, the algorithm will only use the adjusted training data if it has potential to improve the prediction accuracy. Otherwise, it will consider actual training data (without any adjustment) to build the model.

## 2 Methodology

### 2.1 Detection Methods

The three detection methods ‘C1’, ‘C2’ and ‘C3’ are used in the early aberration reporting system (EARS), the syndromic surveillance system of Centers for Disease Control and Prevention (CDC), USA to detect deviations in present data compared to the historic mean [9, 10]. These methods are also used in the CDC’s BioSense system to combat ‘health situational awareness’ and ‘event recognition and response’ [10]. They are also referred as cumulative sum (CUSUM) methods [11, 12]. In this work, we use these three methods to determine false drop or jump days in the training data. Once confirmed, each drop or jump day’s reported infected COVID-19 cases will be replaced by a adjusted value. Then the training data for any model will be based on this adjusted data.

Let *Y* (*t*) denotes the number of confirmed infected cases on the *t*^*th*^ day, then the C1 statistic refers to

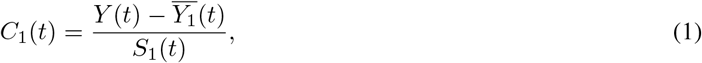

where, 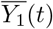 is the moving average and *S*_1_(*t*) is the standard deviation of the number of infected cases since the past seven days. They can be calculated as

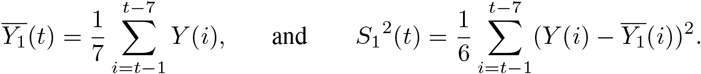

The sample average and the sample standard deviation used in the C1 are based on the 7 days prior to the current observation. Therefore, the C1 indicates the normalized value of the present day infected cases (*Y* (*t*)) considering the last 7 days average infected cases 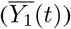 and the corresponding variation 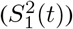. In EARS system, *C*_1_(*t*) *>* 3 indicates a signals at time *t*. In other words, a signal means the *Y* (*t*) exceeds the three sample standard deviations higher than the sample mean. Note that, the EARS only concerns about the higher reporting cases. However, in this work, we are interested in both drop and jump. Therefore, in this context, *C*_1_(*t*) *>* 3 denotes a possible jump and *C*_1_(*t*) *<* −3 refers to a possible drop. The word, “possible” means a drop (or jump) could be a natural trend or a false drop (or false jump). We are only interested in detecting false drops or false jumps.

The C2 statistic is similar to the C1 but it also includes a 1-day lag while calculating the moving average and standard deviation. That is

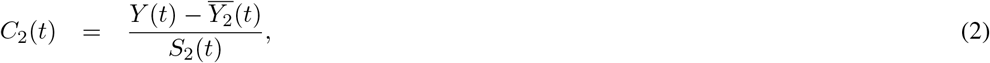

where, 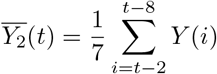, and 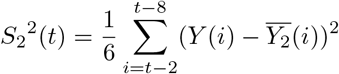.

Note that, [10] defined the C2 statistic with a 2-day lag. However, we observe better results (empirically) if a lag of 1-day is considered instead of 2 days. Hence, we have considered a 1-day lag in the C2 statistic. Similar to the C1 statistic, if |*C*_2_(*t*)| *>* 3, we will check the next subsequent days for a possible jump or drop.

The C3 statistic for the *t*^*th*^ day uses the C2 statistics for the *t*^*th*^ day and the previous two days, as

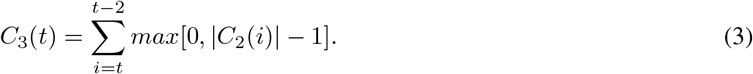

We conclude there is a possible jump or drop when *C*_3_(*t*) *>* 2. In EARS, the expression of *C*_3_(*t*) has *C*_2_(*i*) instead of its absolute value as the objective is to check a jump (increase in infected reported cases) not a drop [10].

### 2.2 Adjustment

The detection methods described in the previous section are used to detect any jump or drop in the training data. This section distinguishes between the actual trend and false drop/jump in the reported infected cases. In identifying a jump or drop, we constrain the length of a jump or drop to be categorized as sharp. The idea behind this is that if the trend of jump or drop continues for an extended period (number of days), it would be considered an actual trend. Hence, we do not require any adjustment in the existing training data to eliminate its impact. Based on emperical results, we have set this upper bound for the duration of a jump or drop to be five days. In other words, if that extended period is more than five days, then we consider it an actual trend.

Once detection method confirms a possible jump or drop for *i*^*th*^ day, we check the same for subsequent five days, and identify the duration of the jump or drop as *j* − *i*, where *j*^*th*^ day is the first day after *i*^*th*^ day without a drop or jump. Then, we propose using the mean of the moving average (*Y*_1_(*i*): as defined in the previous section) of the number of daily confirmed cases in the last seven days and the number of infected cases just after the jump or drop (*Y* (*j* + 1)), to adjust a sharp jump or drop. If the number of daily confirmed cases is outside the specific cut-offs (with respect to C-statistics) from the *i*^*th*^ day to the *j*^*th*^ day, and j-i ≤ 5, then *Y* (*t*) for t ∈ [i, j] would be replaced by

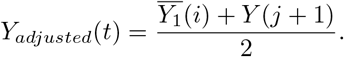

After the adjustment, we consider the root mean square error (RMSE) calculated for the assessment period and compare the values before and after adjusting. If the adjustment has resulted in a decreased RMSE value for the assessment period, then we predict using the adjusted data or else use the original prediction.

Algorithm 1 shows automatic detection and adjustment of the jumps and drops in the training data using the C1, C2 or C3 detection methods.

#### Algorithm 1 Algorithm to identify and adjust jumps and drops

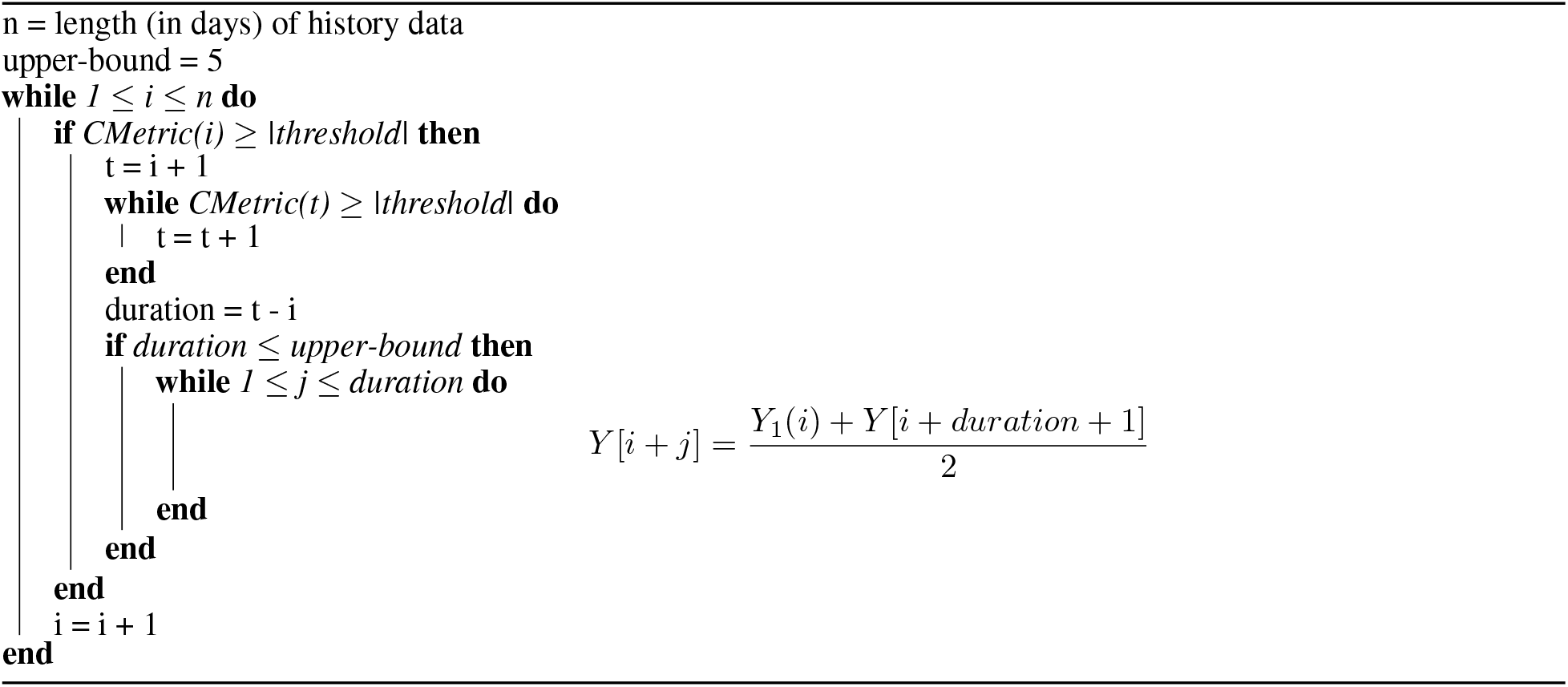

## 3 Jump-Drop Adjusted Prediction using Modified SIS Model

In this section, we show the use of jump-drop adjusted training data for prediction using a recently proposed modified Susceptible-Infected-Susceptible (SIS) [8]. However, proposed jump-drop adjustment algorithm can be applied to a training data required for any epidemiological model to improve the prediction accuracy. The main motivation to consider an SIS model for COVID-19 prediction because it considers re-infection into the model. As it is evident from several studies that re-infection is quite common in COVID-19 [13, 14, 15, 16].

The modified SIS model determines the cumulative number of infected cases (COVID-19) at a specific time point. [8] developed a dynamic data-driven algorithm that estimates the model parameters based on an optimally chosen training phase for prediction of the cumulative infected cases. The model also accounts for deaths due to disease. Their estimation process is useful when the disease under study (like COVID-19) switches its spreading pattern over time. The following equations can describe the modified SIS model proposed by [8],

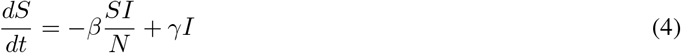

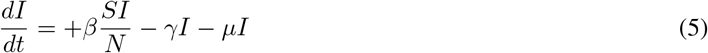

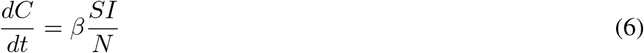

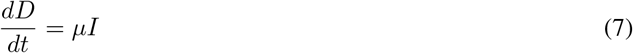

with,

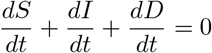

Here t denotes the time where the smallest unit is a day. S and I denote the susceptible and the infected number of people in the population, respectively. C denotes the cumulative infected cases (re-infection is also a count) and D denotes the total deaths caused by the disease. We assume that the death rate from any other cause is the same as the birth rate. The total population size is N. The parameter *β* denotes the average number of individuals infected per day from an infected person [17]. In other words, it is the contact rates between infected and susceptible [18]. *γ* stands for the recovery rate.

Equation (4) represents the rate of change in the population of susceptible compartment. Here, 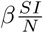 is the number of people get infected daily and are removed from the *S* compartment. The *γ*I is the number of infected individuals recover from the illness daily and are added back to the *S* compartment. The recovery rate, *γ* is assumed to be 1/T, where T is the average duration for which infection lasts in an infected person [2, 19]. Equations (5) and (6) represent the rate of change in population of the infected compartment and the cumulative number of cases, respectively. *µ* is the mortality rate of the infection and daily *µ*I is the number of fatal infected cases, as shown in equation (7).

As argued by [8], the SIS model parameters are generally constant for the entire duration of the study period. However, since the contact rate can change a lot over time in the case of COVID-19, it would be more appropriate to make predictions based on a shorter training phase. Hence, based on the historical data, the training phase has been dynamically chosen. As illustrated in Figure 2, the study period is divided in two parts, the training and the prediction phase. The current date is denoted by *T*_*Current*_. The minimum length of the training period is *T*_*Start*_ and the corresponding training interval is [*T*_*Current*_ − *T*_*Start*_ + 1, *T*_*Current*_]. For selecting the optimal training phase, at most *T*_*Limit*_ number of days (set by the user) can be added to the minimum training period. Therefore, the training period is [*T*_*Current*_ − *T*_*Start*_ + 1 − *t, T*_*Current*_], where 0 ≤ *t* ≤ *T*_*Limit*_. *T*_*Pred*_ denotes the length of the prediction phase.

**Figure 2:**
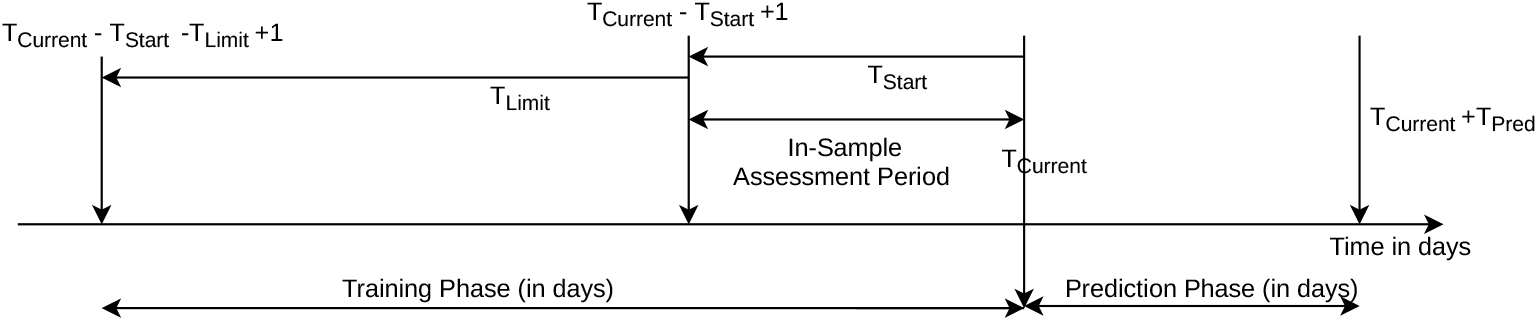
Partition of the study period into different training and prediction time points [8]

Here, the objective is to optimally choose a training phase so that we can accurately predict the near future using proper model parameters. To compare different models, we use the minimum training phase as the fixed in-sample assessment period. The root mean square error (RMSE) has been used as an criteria to choose the optimal training period,

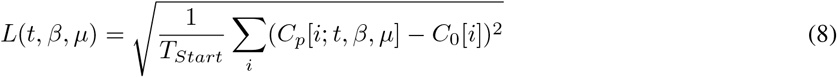

where *T*_*Current*_ - *T*_*Start*_ + 1 ⩽ *i* ⩽ *T*_*Current*_. The observed cumulative infected cases on the *i*^*th*^ day is given by *C*_*o*_[*i*] and *C*_*p*_[*i*; *t, β, µ*] denotes the predicted cumulative infected cases on *i*^*th*^ day from modified SIS model. The *t*_*opt*_ is the value of *t* for which (8) is minimized. Similarly, 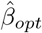 and 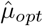 are the values of 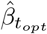 and 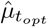, respectively. The assessment of prediction by a model is done by using (8) but replacing *T*_*Start*_, *t, β* and *µ*, by *T*_*Pred*_, *t*_*opt*_, 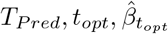 and 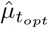, respectively; where 0 ≤ *i* ≤ *T*_*Pred*_. The recovery period in India from COVID-19 infection is on an average 14 days [3]. *γ* can thus be calculated as 1/(duration for recovery), that is, 1/14.

## 4 PREDICTION OF CUMULATIVE INFECTED CASES

### 4.1 R-Package

For easy use of the jump-drop adjusted prediction model, an R package has been developed and is available at https://github.com/RashiMohta/COVID-19-cases-prediction. Given the daily infected COVID-19 data (say for a state) and other input parameter values, the R-package would predict the cumulative number of cases using both the original data and the data adjusted using the given bound metric and return the prediction with lesser root mean squared error in the in-sample assessment period.

### 4.2 Predicting the cumulative number of COVID-19 cases

To demonstrate the performance of the proposed jump-drop adjustment algorithm, we use the developed R-package to predict the cumulative infected cases in the capital of India, Delhi. The data containing the number of daily confirmed cases of COVID-19 is publicly available on https://www.covid19india.org/. In Table 1, we consider six different *T*_*current*_ as 12 May 2020 (in (A)), 12 June 2020 (in (B)), 12 July 2020 (in (C)), 14 September 2020 (in (D)), 12 April 2021 (in (E)) and 27 April 2021 (in (F)), to observe the impact of adjusting the jump drops using the proposed algorithm. We have set the upper limit (*T*_*Limit*_) of the number of additional days that can be added to the minimum training period-as 30. The prediction has been made using the original and the jump-drop adjusted data for the period *T*_*Current*_ + 1 to 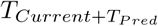 where *T*_*Pred*_ has been taken as 20 days. Table 1 shows the optimal values of *µ* and *β* calculated for different prediction periods and the corresponding RMSE values for the assessment and the prediction period. Note that, in all the scenarios, we have used modified SIS model described in Section 3.

**Table 1:** Using the data from Delhi, the estimated optimal parameters of the modified SIS model, RMSE of assessment and prediction periods with original training data and with adjusted training data using C1, C2, C3 statistics. In all the six scenarios, the assessment and prediction periods are taken as 20 days.

| Graphs No.<br>in Figure 3 | $T_{Current}$ | Variable parameter | Original | With adjustment | | |
| --- | --- | --- | --- | --- | --- | --- |
|  |  |  |  | C1 | C2 | C3 |
| (A) | 12 May<br>2020 | $\hat{\mu}_{t_{opt}}$ | 0.063 | 0.053 | 0.048 | 0.06 |
| | | $\hat{\beta}_{t_{opt}}$ | 0.19 | 0.19 | 0.19 | 0.18 |
|  |  | Opt. training period | 41 | 40 | 41 | 35 |
|  |  | RMSE (Assessment) | 296.70 | 316.10 | 311.60 | **248.02** |
|  |  | RMSE (Prediction) | 10984.22 | 8870.75 | 7395.23 | **1171.24** |
| (B) | 12 Jun<br>2020 | $\hat{\mu}_{t_{opt}}$ | 0.002 | 0.018 | 0.001 | 0.008 |
| | | $\hat{\beta}_{t_{opt}}$ | 0.12 | 0.13 | 0.12 | 0.13 |
|  |  | Opt. training period | 20 | 20 | 20 | 24 |
|  |  | RMSE (Assessment) | 1417.65 | 1374.42 | 1291.85 | **1148.56** |
|  |  | RMSE (Prediction) | 12654.64 | 11648.56 | 8285.07 | **5105.93** |
| (C) | 12 Jul<br>2020 | $\hat{\mu}_{t_{opt}}$ | 0.097 | 0.095 | 0.099 | 0.098 |
| | | $\hat{\beta}_{t_{opt}}$ | 0.14 | 0.14 | 0.14 | 0.15 |
|  |  | Opt. training period | 27 | 27 | 26 | 24 |
|  |  | RMSE (Assessment) | 2649.52 | 2430.44 | **2193.65** | 2381.78 |
|  |  | RMSE (Prediction) | 11092.92 | 10206.85 | **7865.84** | 12247.35 |
| (D) | 14 Sep<br>2020 | $\hat{\mu}_{t_{opt}}$ | 0.013 | 0.013 | 0.013 | 0.001 |
| | | $\hat{\beta}_{t_{opt}}$ | 0.14 | 0.14 | 0.14 | 0.13 |
|  |  | Opt. training period | 20 | 20 | 20 | 22 |
|  |  | RMSE (Assessment) | 3350.79 | 3350.79 | 3350.79 | **2886.15** |
|  |  | RMSE (Prediction) | 31161.71 | 31161.71 | 31161.71 | **25972.73** |
| (E) | 12 Apr<br>2021 | $\hat{\mu}_{t_{opt}}$ | 0.008 | 0.006 | 0.007 | 0.069 |
| | | $\hat{\beta}_{t_{opt}}$ | 0.22 | 0.22 | 0.22 | 0.28 |
|  |  | Opt. training period | 20 | 20 | 20 | 20 |
|  |  | RMSE (Assessment) | 6140.81 | 6162.93 | 6367.32 | **5374.55** |
|  |  | RMSE (Prediction) | 196356.66 | 101354.75 | 92301.01 | **73298.98** |
| (F) | 27 Apr<br>2021 | $\hat{\mu}_{t_{opt}}$ | 0.089 | 0.089 | 0.089 | 0.069 |
| | | $\hat{\beta}_{t_{opt}}$ | 0.26 | 0.26 | 0.26 | 0.24 |
|  |  | Opt. training period | 49 | 49 | 49 | 49 |
|  |  | RMSE (Assessment) | 25354.74 | 25354.74 | 26088.72 | **23272.09** |
|  |  | RMSE (Prediction) | 443016.61 | 443016.61 | 436018.05 | **162015.09** |

In practice, when we do prediction for future, we do not have the observe infected cases for the prediction period. Therefore, as discussed earlier, the RSME of the in-sample assessment period is used to chose between the with or without adjustment prediction and the corresponding adjustment method (C1, C2 or C3). From Table 1, we observe that the five out of the six scenarios the adjustment with C3 is giving the best results. Notice that, there is an increasing trend in RMSE values over the time (calendar days). It is due to prediction of cumulative infected cases, which always increase over the time. For example, in the first scenario (A), the cumulative infected cases in the training period was less than 10,000. Thus, corresponding RMSE values (for C3, RMSE-assessment: 248.02 and RMSE-prediction: 1171.24) are expected to be less. Parameters 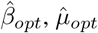 and the optimal training period are also changing for different scenarios justifying the use of dynamic data-driven modified SIS model. Figure 3 shows the graphical comparison of the scenarios presented in Table 1. In Figure 3, all the cases, the observed curve and adjusted prediction curve are close to each other. Table 2 (in Appendix) shows the prediction for Maharashtra and Madhya Pradesh, the two other Indian states.

**Table 2:** Using the data from Maharashtra and Madhya Pradesh, the estimated optimal parameters of the modified SIS model, RMSE of assessment and prediction periods with original training data and with adjusted training data using C1, C2, C3 statistics. In all the seven scenarios, the assessment and prediction periods are taken as 20 days.

| State | $T_{Current}$ | Variable parameter | Original | With adjustment | | |
| --- | --- | --- | --- | --- | --- | --- |
|  |  |  |  | C1 | C2 | C3 |
| Maharashtra | 12 May 2020 | $\hat{\mu}_{t_{opt}}$ | 0.096 | 0.036 | 0.095 | 0.033 |
| | | $\hat{\beta}_{t_{opt}}$ | 0.23 | 0.16 | 0.22 | 0.16 |
|  |  | Opt. training period | 36 | 28 | 32 | 25 |
|  |  | RMSE (Assessment) | 1012.60 | 971.56 | 955.99 | **945.98** |
|  |  | RMSE (Prediction) | 56252.22 | 20378.43 | 38767.01 | **18147.96** |
| Maharashtra | 21 Jun 2020 | $\hat{\mu}_{t_{opt}}$ | 0.004 | 0.005 | 0.004 | 0.001 |
| | | $\hat{\beta}_{t_{opt}}$ | 0.09 | 0.09 | 0.09 | 0.08 |
|  |  | Opt. training period | 27 | 25 | 25 | 21 |
|  |  | RMSE (Assessment) | 9155.62 | 9545.89 | 9435.10 | **8726.17** |
|  |  | RMSE (Prediction) | 41659.11 | 37311.09 | 37230.61 | **30591.41** |
| Maharashtra | 19 Sep 2020 | $\hat{\mu}_{t_{opt}}$ | 0.018 | 0.018 | 0.018 | 0.001 |
| | | $\hat{\beta}_{t_{opt}}$ | 0.10 | 0.10 | 0.10 | 0.09 |
|  |  | Opt. training period | 20 | 20 | 20 | 21 |
|  |  | RMSE (Assessment) | 23770.72 | 23770.72 | 23770.72 | **21489.21** |
|  |  | RMSE (Prediction) | 171609.02 | 171609.02 | 171609.02 | **134484.32** |
| Maharashtra | 14 Oct 2020 | $\hat{\mu}_{t_{opt}}$ | 0.027 | 0.027 | 0.027 | 0.031 |
| | | $\hat{\beta}_{t_{opt}}$ | 0.07 | 0.07 | 0.07 | 0.07 |
|  |  | Opt. training period | 25 | 25 | 25 | 21 |
|  |  | RMSE (Assessment) | 13876.04 | 13876.04 | 13876.04 | **13741.02** |
|  |  | RMSE (Prediction) | 43017.37 | 43017.37 | 43017.37 | **29206.76** |
| Maharashtra | 17 Apr 2021 | $\hat{\mu}_{t_{opt}}$ | 0.069 | 0.069 | 0.069 | 0.043 |
| | | $\hat{\beta}_{t_{opt}}$ | 0.18 | 0.18 | 0.18 | 0.15 |
|  |  | Opt. training period | 33 | 33 | 33 | 25 |
|  |  | RMSE (Assessment) | 57767.96 | 57767.96 | 57767.96 | **57094.2** |
|  |  | RMSE (Prediction) | 884749.37 | 884749.37 | 884749.37 | **525393.53** |
| Madhya Pradesh | 19 Oct 2020 | $\hat{\mu}_{t_{opt}}$ | 0.089 | 0.055 | 0.055 | 0.056 |
| | | $\hat{\beta}_{t_{opt}}$ | 0.15 | 0.10 | 0.10 | 0.10 |
|  |  | Opt. training period | 43 | 26 | 25 | 22 |
|  |  | RMSE (Assessment) | 2141.95 | **1521.49** | 1545.69 | 1597.72 |
|  |  | RMSE (Prediction) | 11708.52 | **5065.79** | 5051.13 | 2525.85 |
| Madhya Pradesh | 15 Aug 2021 | $\hat{\mu}_{t_{opt}}$ | 0.085 | 0.003 | 0.003 | 0.002 |
| | | $\hat{\beta}_{t_{opt}}$ | 0.13 | 0.05 | 0.05 | 0.05 |
|  |  | Opt. training period | 20 | 48 | 48 | 48 |
|  |  | RMSE (Assessment) | **16.20** | 21.55 | 20.81 | 18.92 |
|  |  | RMSE (Prediction) | **29.74** | 2066.62 | 2065.15 | 2049.2 |
| Madhya Pradesh | 12 May 2020 | $\hat{\mu}_{t_{opt}}$ | 0.040 | 0.063 | 0.083 | 0.087 |
| | | $\hat{\beta}_{t_{opt}}$ | 0.12 | 0.15 | 0.17 | 0.20 |
|  |  | Opt. training period | 28 | 28 | 28 | 40 |
|  |  | RMSE (Assessment) | 129.24 | 139.67 | 142.37 | **88.62** |
|  |  | RMSE (Prediction) | 316.52 | **307.81** | 430.65 | 3126 |

**Figure 3:**
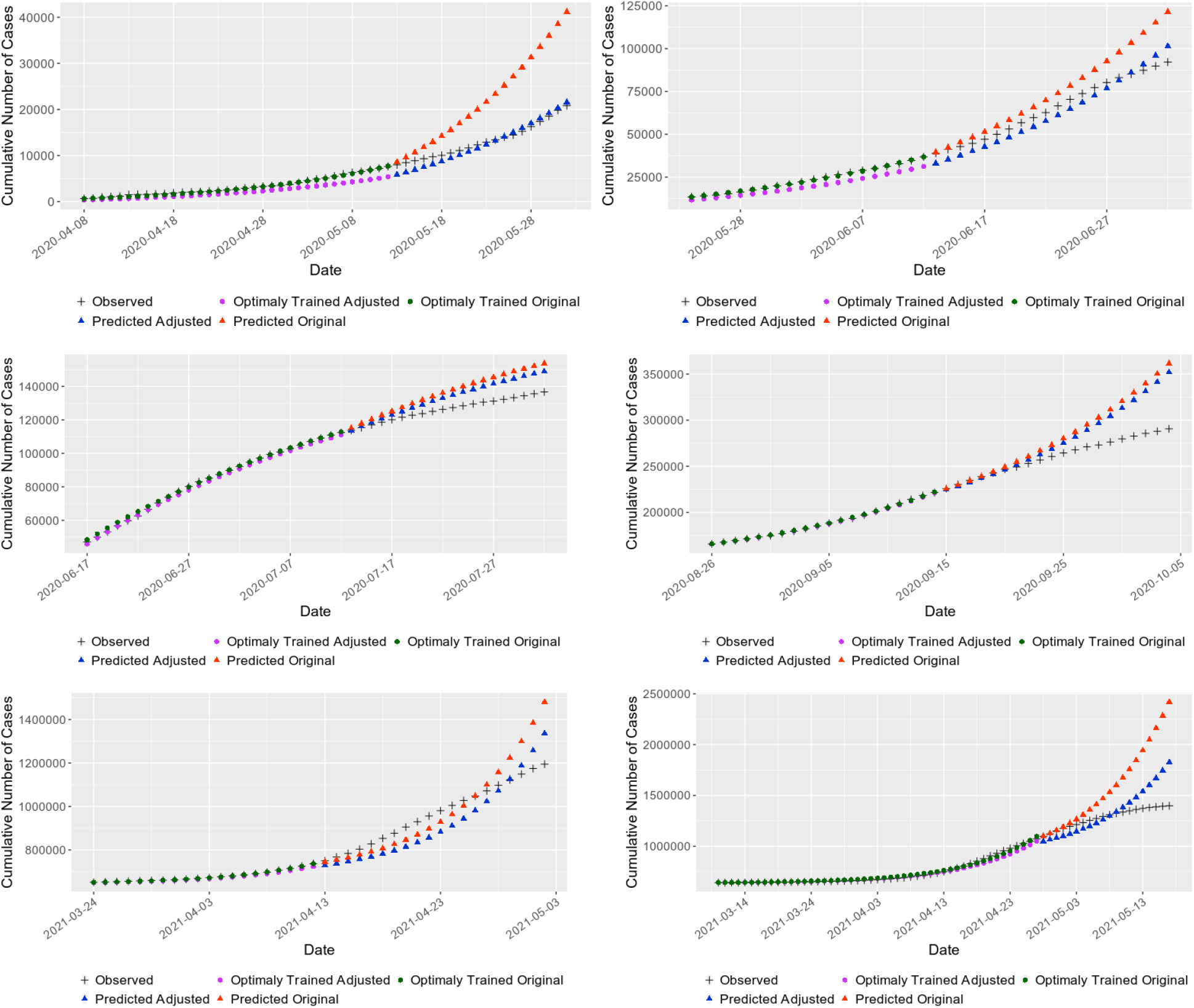
Predictions made with and without adjustments for Delhi at different periods using best performing metric.

## 5 Conclusion

This work established the importance of adjusting the sharp (false) jumps and drops observed in infected COVID-19 cases while making the prediction. The proposed algorithm uses three detection methods, C1, C2, and C3, to automatically detect these jumps and drops and adjust the corresponding values. We then made predictions using all three methods and compared the results with the same model without making these adjustments. The RMSE values for the considered dates supported the assumption that a decrease in RMSE in the assessment period should imply better accuracy in the prediction. Looking at the RMSE values for the prediction period, we observed a significant increase in accuracy.

For a specific date (current day), the proposed algorithm will also check whether any adjustment is required or not based on the ‘in sample assessment period.’ If so, the prediction will be made using adjusted training data. Otherwise, the algorithm will use original training data (without adjustment). In other words, the proposed algorithm is adaptive enough to automatically choose the right training data with or without adjustment. However, RMSE in the assessment period may be the lowest for one detection method (say C3), but the RMSE in the prediction period (when observed data is available for this period) is lowest for another detection method (say C1). In the last scenario of Table 2 (Appendix), we observe this phenomena. It indicates the infection trend during the prediction period has changed substantially from that of the training period.

## Data Availability

The data used in this work is publicly available on https://www.covid19india.org/

## Data Availability Statement

The data used in this work is publicly available on https://www.covid19india.org/.

## Acknowledgments

Rashi Mohta, Prathapani Sravya would like to acknowledge Samsung Fellowship for this work. Palash Ghosh would like to acknowledge support by ICMR Centre for Excellence, Grant no. 5/3/8/20/2019-ITR.

## Appendix

## Notes

### Competing Interest Statement

The authors have declared no competing interest.

### Author Declarations

https://www.covid19india.org/

